# Caregiver Experiences and Perceptions of Managing Sundowning Behaviour in Dementia: A Protocol for a Qualitative Evidence Synthesis

**DOI:** 10.1101/2025.10.14.25338031

**Authors:** Ravi Shankar, Jun Wen Joshua, Xu Qian

## Abstract

**Background:** Sundowning syndrome, characterized by increased confusion, agitation, and behavioral disturbances during late afternoon and evening hours, affects up to 66% of individuals with dementia. This phenomenon significantly impacts caregivers who must manage these challenging behaviors while experiencing physical and emotional exhaustion. Despite its prevalence, caregiver perspectives on managing sundowning remain poorly synthesized in the literature.

**Objective:** To systematically identify, appraise, and synthesize qualitative evidence on caregiver experiences and perceptions of managing sundowning behavior in individuals with dementia across diverse care settings.

**Methods:** This qualitative evidence synthesis will follow the Enhancing Transparency in Reporting the Synthesis of Qualitative Research (ENTREQ) guidelines and Joanna Briggs Institute methodology for qualitative systematic reviews. Comprehensive searches will be conducted in MEDLINE, CINAHL, PsycINFO, EMBASE, Scopus, and Web of Science from inception to December 2025. Two reviewers will independently screen studies using Covidence software, extract data, and assess quality using the Critical Appraisal Skills Programme (CASP) qualitative checklist. Data synthesis will employ meta-aggregation and thematic synthesis approaches.

**Data Synthesis:** The synthesis will use Noblit and Hare’s meta-ethnographic approach combined with Thomas and Harden’s thematic synthesis. Line-by-line coding will identify descriptive themes, which will be developed into analytical themes. The Confidence in the Evidence from Reviews of Qualitative research (GRADE-CERQual) approach will assess confidence in review findings.

## Introduction

Sundowning syndrome, also known as sundown syndrome or late-day confusion, represents one of the most challenging neuropsychiatric phenomena in dementia care, affecting between 2.5% to 66% of individuals with dementia depending on the diagnostic criteria and assessment methods used [1-8]. This syndrome manifests as a marked increase in confusion, agitation, anxiety, pacing, wandering, and other behavioral disturbances that typically emerge or intensify during late afternoon and evening hours, creating significant management challenges for caregivers across all care settings [9]. The temporal pattern of sundowning aligns with circadian rhythm disruptions common in dementia, though the exact pathophysiology remains incompletely understood, with proposed mechanisms including dysfunction of the suprachiasmatic nucleus, melatonin dysregulation, environmental factors, and unmet physical or psychological needs [10].

The impact of sundowning on caregivers is profound and multifaceted, encompassing physical exhaustion from managing disruptive behaviors during evening hours when they themselves are fatigued, emotional distress from witnessing their loved one’s confusion and agitation, disrupted sleep patterns leading to chronic sleep deprivation, and increased risk of caregiver burnout and depression [11, 12]. Family caregivers, who provide approximately 80% of dementia care in community settings, often report that sundowning behaviors are among the most distressing aspects of dementia caregiving and frequently precipitate decisions for institutional placement [13]. Professional caregivers in long-term care facilities similarly identify sundowning as a significant challenge, particularly during shift changes when staffing levels may be reduced and multiple residents may simultaneously experience increased agitation [14].

Recent research has begun addressing multiple dimensions of caregiver well-being, including the effectiveness of technology-based interventions for reducing caregiver burden, the development and validation of caregiver burden assessment protocols, and psychosocial interventions to enhance caregiver well-being across various chronic illness populations. While these studies contribute to understanding caregiver burden measurement and intervention effectiveness [15-19], sundowning-specific caregiver experiences remain underexplored in systematic qualitative syntheses that could inform targeted support strategies.

Current evidence regarding sundowning management primarily focuses on pharmacological and non-pharmacological interventions from clinical perspectives, with limited synthesis of caregiver experiences and perceptions of managing these behaviors in real-world settings [1]. Existing systematic reviews have examined the effectiveness of light therapy, melatonin supplementation, structured activity programs, and environmental modifications, but these reviews predominantly synthesize quantitative outcome data without capturing the nuanced experiences of caregivers who implement these strategies daily [20]. The qualitative dimensions of sundowning management, including how caregivers recognize early signs, adapt strategies to individual needs, navigate emotional challenges, and balance competing demands, remain fragmented across individual studies without comprehensive synthesis.

The caregiving context for sundowning management varies significantly across settings and relationships, with family caregivers in home settings facing different challenges than professional caregivers in institutional environments [21]. Cultural factors also influence caregiver perceptions and management approaches, with varying beliefs about the causes of behavioral symptoms and acceptable intervention strategies across different cultural groups [22]. Additionally, the availability of resources, support systems, and respite care significantly impacts caregiver capacity to manage sundowning behaviors effectively, yet these contextual factors are rarely examined comprehensively in existing literature reviews.

Recent qualitative studies have begun exploring caregiver perspectives on sundowning management, revealing complex emotional responses, adaptive strategies, and support needs that quantitative measures alone cannot capture [23]. Caregivers report developing intuitive knowledge about triggers and patterns specific to their care recipient, implementing creative environmental modifications, and struggling with decisions about medication use versus behavioral interventions [24]. These rich qualitative insights remain scattered across individual studies, limiting their potential to inform evidence-based practice guidelines and caregiver support programs. A systematic synthesis of this qualitative evidence is urgently needed to develop comprehensive understanding of caregiver experiences and inform person-centered approaches to sundowning management.

### Theoretical Framework

This qualitative evidence synthesis will be guided by the Stress Process Model for caregiving, originally developed by Pearlin et al. [25] and adapted for dementia caregiving contexts. This framework conceptualizes caregiving as a complex process involving multiple interrelated components that influence caregiver outcomes and adaptation strategies. The model distinguishes between primary stressors directly related to caregiving tasks and care recipient symptoms, secondary stressors arising from role conflicts and lifestyle disruptions, mediating factors including coping strategies and social support, and outcomes encompassing physical and psychological well-being [25].

Within this framework, sundowning behaviors represent a primary stressor that directly challenges caregiver resources through their unpredictable timing, intensity, and resistance to intervention. The model acknowledges that caregiver appraisal of these behaviors significantly influences their stress response and choice of management strategies. Caregivers who perceive sundowning as an inevitable aspect of dementia may adopt different coping approaches than those who view it as a modifiable symptom requiring active intervention [26]. The framework also recognizes the role of contextual factors, including caregiving setting, relationship quality, cultural background, and available resources, in shaping caregiver experiences and responses to sundowning behaviors.

Secondary stressors in the context of sundowning management include role strain from balancing caregiving with other responsibilities during evening hours traditionally reserved for family time or rest, social isolation resulting from inability to engage in evening activities, and family conflicts about appropriate management approaches [27]. The framework’s attention to stress proliferation helps explain how sundowning-related challenges can cascade into broader life disruptions, affecting employment, relationships, and caregiver identity. This comprehensive view of stress processes will guide data extraction and synthesis, ensuring attention to both direct experiences of managing sundowning and broader impacts on caregiver well-being.

The model’s emphasis on mediating factors provides a lens for examining how caregivers develop and deploy coping strategies specific to sundowning management. These may include problem-focused strategies such as environmental modifications and activity scheduling, emotion-focused approaches including acceptance and meaning-making, and social coping through support group participation or respite care utilization [28]. The framework recognizes that effective coping is context-dependent, with strategies that work in one situation potentially proving ineffective or counterproductive in another. This nuanced understanding of coping processes will inform the synthesis of diverse caregiver experiences across different contexts and care relationships.

### Objectives

The primary objective of this qualitative evidence synthesis is to systematically identify, critically appraise, and synthesize available qualitative evidence regarding caregiver experiences and perceptions of managing sundowning behavior in individuals with dementia, encompassing both family and professional caregivers across diverse care settings including home, residential, and institutional environments. This synthesis aims to develop a comprehensive understanding of how caregivers recognize, interpret, respond to, and cope with sundowning behaviors, examining the emotional, practical, and relational dimensions of this caregiving challenge while identifying patterns in caregiver experiences that can inform evidence-based support interventions and practice guidelines. Specific objectives include synthesizing caregiver descriptions of sundowning manifestations and their impact on daily routines and well-being; exploring the strategies caregivers develop and implement to prevent or manage sundowning episodes, including their perceived effectiveness and contextual factors influencing strategy selection; examining caregiver emotional responses, meaning-making processes, and adaptation over time; identifying support needs, resource gaps, and facilitators of effective sundowning management from caregiver perspectives; and exploring variations in experiences across different caregiver types, care settings, and cultural contexts to understand how these factors shape management approaches and outcomes.

## Methods

### Study Design

This qualitative evidence synthesis will follow a systematic approach to identifying, appraising, and synthesizing qualitative research on caregiver experiences of managing sundowning in dementia. The protocol adheres to the Enhancing Transparency in Reporting the Synthesis of Qualitative Research (ENTREQ) statement to ensure comprehensive reporting [29]. The Joanna Briggs Institute (JBI) methodology for qualitative systematic reviews will guide the review process, providing a rigorous framework for aggregating qualitative findings while preserving the richness and complexity of caregiver experiences [30]. The synthesis will employ both meta-aggregation and thematic synthesis approaches to develop comprehensive understanding of the phenomenon while maintaining connection to primary study findings.

### Search Strategy

A comprehensive search strategy has been developed in consultation with a health sciences librarian experienced in systematic review methodology. The strategy combines controlled vocabulary terms specific to each database with free-text keywords to ensure maximum sensitivity. The search will encompass multiple electronic databases from their inception to December 2025, including MEDLINE via PubMed, CINAHL Complete, PsycINFO via APA PsycNet, EMBASE via Elsevier, Scopus, and Web of Science Core Collection. These databases were selected to ensure coverage of medical, nursing, psychological, and interdisciplinary literature relevant to dementia caregiving.

The search strategy employs three main concept blocks combined with Boolean operators. The first block captures dementia and related conditions using terms including “dementia,” “Alzheimer’s disease,” “cognitive impairment,” “neurocognitive disorder,” and related MeSH/subject headings. The second block targets sundowning phenomena through terms such as “sundowning,” “sundown syndrome,” “evening agitation,” “late-day confusion,” “circadian rhythm disturbance,” and “behavioral and psychological symptoms of dementia” with temporal qualifiers. The third block focuses on caregivers and their experiences using terms including “caregiver,” “carer,” “family,” “spouse,” “adult children,” “care staff,” “experience,” “perception,” “attitude,” “coping,” and “management.” No language or date restrictions will be applied initially, with translation services utilized for potentially relevant non-English articles.

Grey literature searches will include ProQuest Dissertations and Theses Global, conference proceedings from major dementia and gerontology conferences, and websites of relevant organizations including the Alzheimer’s Association, Alzheimer’s Disease International, and national dementia organizations. Reference lists of included studies and relevant systematic reviews will be hand-searched for additional sources. Authors of included studies will be contacted for unpublished or in-progress research when appropriate. Forward citation searching will be conducted for key papers using Google Scholar and Web of Science.

### Eligibility Criteria

Studies will be included if they employ qualitative research methods including phenomenology, grounded theory, ethnography, descriptive qualitative approaches, or mixed-methods studies with substantial qualitative components that can be extracted separately. The population of interest encompasses all types of caregivers including family members, friends, professional care staff, and healthcare providers who have direct experience managing sundowning behaviors in individuals with any type or stage of dementia. The phenomenon of interest is caregiver experiences, perceptions, attitudes, and practices related to recognizing, preventing, managing, or coping with sundowning behaviors, including emotional responses, decision-making processes, strategy development, and support needs.

Studies will be included regardless of care setting, encompassing home care, adult day programs, assisted living facilities, nursing homes, memory care units, and hospital settings. Both published and unpublished studies will be considered, including peer-reviewed articles, dissertations, theses, and research reports. Studies will be excluded if they focus exclusively on quantitative outcomes without qualitative exploration of experiences, present only researcher interpretations without participant voices, focus on behavioral symptoms without specific attention to sundowning or temporal patterns, or examine only care recipient perspectives without caregiver input. Conference abstracts without full reports, opinion pieces, and clinical guidelines without empirical qualitative data will also be excluded.

### Study Selection

All identified citations will be imported into Covidence systematic review software for screening and management. After automated and manual deduplication, two reviewers will independently screen titles and abstracts against the eligibility criteria using standardized screening forms. The screening form will be piloted on 25 citations to ensure consistency between reviewers. Disagreements will be resolved through discussion, with a third reviewer consulted when consensus cannot be reached. Full texts of potentially eligible studies will be obtained and independently assessed by two reviewers using detailed eligibility criteria. Reasons for exclusion at the full-text stage will be documented and reported in a PRISMA flow diagram.

### Data Extraction

A standardized data extraction form will be developed and piloted on three included studies before full implementation. Two reviewers will independently extract data from all included studies, with discrepancies resolved through discussion. Extracted data will include study characteristics (authors, year, country, setting, methodology, theoretical framework), participant characteristics (caregiver type, relationship to care recipient, demographics, caregiving duration), care recipient characteristics (dementia type and stage, sundowning severity), study findings (themes, categories, metaphors, concepts), and supporting participant quotes that illustrate key findings.

Particular attention will be paid to extracting rich descriptive data about caregiver experiences, including emotional responses to sundowning, specific management strategies employed, perceived effectiveness of different approaches, factors influencing strategy selection, support needs and resource utilization, and impacts on caregiver well-being and caregiving decisions. Contextual information about care settings, cultural factors, and health system characteristics will also be extracted to facilitate assessment of transferability.

### Quality Assessment

Methodological quality will be assessed using the Critical Appraisal Skills Programme (CASP) Qualitative Checklist, which evaluates ten key aspects of qualitative research including research aims, methodology appropriateness, research design, recruitment strategy, data collection, researcher reflexivity, ethical considerations, data analysis rigor, findings clarity, and research value [31]. Two reviewers will independently assess each study, with disagreements resolved through discussion. Studies will not be excluded based on quality assessment; rather, quality appraisal will inform confidence in individual findings during synthesis and GRADE-CERQual assessment.

The assessment will particularly focus on whether studies provide thick description of caregiver experiences, demonstrate reflexivity regarding researcher influence on data collection and interpretation, show clear links between data and interpretations, and consider diverse perspectives and negative cases. The quality assessment results will be presented in a summary table and narrative description, highlighting methodological strengths and limitations across the body of evidence.

### Data Synthesis

The synthesis will combine Noblit and Hare’s meta-ethnographic approach with Thomas and Harden’s thematic synthesis to develop both interpretive and aggregative understanding of caregiver experiences [32, 33]. The synthesis process will begin with repeated reading of included studies to achieve immersion in the data. Line-by-line coding of findings sections will identify concepts and meanings related to sundowning management experiences. These codes will be compared across studies to identify patterns and relationships, developing descriptive themes that remain close to primary study findings.

The synthesis will then progress to developing analytical themes that go beyond individual studies to generate new interpretive explanations and theoretical insights. This will involve examining relationships between themes, exploring variations and contradictions in caregiver experiences, and considering how contextual factors influence sundowning management. The concept of reciprocal translation will be used to identify common experiences across studies, while refutational synthesis will explore contradictory findings and their potential explanations. A lines-of-argument synthesis will develop overarching interpretations that encompass the full range of caregiver experiences while maintaining groundedness in primary data.

The Confidence in the Evidence from Reviews of Qualitative research (GRADE-CERQual) approach will assess confidence in each review finding across four components: methodological limitations of contributing studies, coherence of the finding, adequacy of data supporting the finding, and relevance of contributing studies to the review question [34]. Each finding will be assigned a confidence rating (high, moderate, low, or very low) with explanations for any downgrading decisions. This assessment will help users understand the strength of evidence supporting each synthesis finding.

## Discussion

This qualitative evidence synthesis will provide the first comprehensive examination of caregiver experiences managing sundowning behaviors in dementia, addressing a critical gap in the literature that has predominantly focused on intervention effectiveness without adequately capturing the lived experiences of those providing daily care. The synthesis findings will illuminate the complex realities of sundowning management from caregiver perspectives, moving beyond clinical descriptions to understand the emotional, practical, and relational dimensions of this caregiving challenge. By synthesizing diverse caregiver voices across different settings and relationships, this review will develop nuanced understanding of how sundowning impacts caregivers and how they develop expertise in recognizing and responding to these challenging behaviors.

The application of the Stress Process Model will enable systematic examination of how sundowning behaviors function as stressors within the broader caregiving context, identifying both direct impacts and cascading effects on caregiver well-being and family dynamics. The synthesis is expected to reveal patterns in how caregivers appraise and respond to sundowning, potentially identifying distinct caregiver typologies based on management approaches, coping styles, and support utilization. These insights will be valuable for developing tailored interventions that align with different caregiver needs and preferences rather than assuming one-size-fits-all approaches to sundowning management.

Expected findings include identification of early warning signs and triggers that experienced caregivers recognize but may not be captured in clinical assessments, creative management strategies developed through trial and error that could benefit other caregivers, emotional and meaning-making processes that help caregivers cope with the distress of witnessing sundowning episodes, and critical support gaps and resource needs from caregiver perspectives. The synthesis may reveal that caregivers develop sophisticated understanding of individual patterns and responses that could inform personalized care approaches. Additionally, the review is likely to identify tensions between recommended evidence-based practices and the realities of implementing these strategies in complex caregiving contexts with limited resources.

The synthesis will likely reveal important variations in experiences based on caregiver characteristics, care settings, and cultural contexts. Family caregivers may describe different challenges than professional caregivers, with unique emotional burdens related to witnessing personality changes in loved ones during sundowning episodes. Cultural factors may influence how caregivers interpret sundowning behaviors, with some viewing them through spiritual or traditional explanatory models that shape management approaches. Understanding these variations will be crucial for developing culturally responsive support programs and ensuring interventions are relevant across diverse caregiving populations.

Methodological strengths of this synthesis include the comprehensive search strategy without language restrictions, rigorous quality assessment using established tools, systematic approach to synthesis combining multiple analytical techniques, and transparent assessment of confidence in findings using GRADE-CERQual. The use of Covidence for study management ensures reproducibility and transparency in the selection process. The combination of meta-ethnographic and thematic synthesis approaches will enable both interpretive depth and systematic aggregation of findings, providing rich understanding while maintaining clear connections to primary data.

Limitations must be acknowledged to appropriately contextualize the synthesis findings. The focus on qualitative evidence means that quantitative data on intervention effectiveness or prevalence will not be included, requiring users to consult complementary quantitative reviews for comprehensive evidence. The quality of synthesis findings will depend on the quality and richness of available primary studies, which may vary considerably. Publication bias may result in underrepresentation of certain caregiver groups or settings if their experiences have not been studied qualitatively. The synthesis of studies across different healthcare systems and time periods may obscure important contextual influences on caregiver experiences.

The implications for practice are substantial, as this synthesis will provide evidence-based understanding of caregiver needs and preferences that can inform support program development, clinical practice guidelines, and health policy. Healthcare providers will gain insights into the caregiver perspective on sundowning that can enhance communication, shared decision-making, and collaborative care planning. The identification of effective caregiver-developed strategies may lead to new intervention approaches that build on experiential expertise. Understanding support gaps and resource needs from caregiver perspectives will help prioritize service development and resource allocation.

For research, this synthesis will identify knowledge gaps requiring future investigation, methodological considerations for studying caregiver experiences of sundowning, and theoretical insights that can guide future research questions and study designs. The synthesis may reveal that certain aspects of sundowning management have received limited qualitative exploration, such as decision-making about medication use, navigation of safety concerns, or long-term adaptation to sundowning patterns. These gaps will provide direction for future qualitative research priorities. The synthesis findings may also suggest the need for intervention studies that incorporate caregiver-identified outcomes and preferences rather than relying solely on clinical measures.

The policy implications include evidence for developing caregiver support policies that address sundowning-specific challenges, insights for improving dementia care standards and quality indicators, and understanding of resource needs for effective sundowning management across care settings. Policymakers will gain appreciation for the complexity of sundowning management and the sophisticated skills caregivers develop, potentially leading to greater recognition and support for caregiver expertise.

The synthesis may highlight systemic barriers to effective sundowning management that require policy intervention, such as inadequate respite care during evening hours or lack of specialized training for professional caregivers.

## Data Availability

All data produced in the present work are contained in the manuscript

